# The secretory IgA (sIgA) response in human milk against the SARS-CoV-2 Spike is highly durable and neutralizing for at least 1 year of lactation post-infection

**DOI:** 10.1101/2023.05.19.23290192

**Authors:** Xiaoqi Yang, Alisa Fox, Claire DeCarlo, Nicole Pineda, Rebecca L.R. Powell

## Abstract

Although in the early pandemic period, COVID-19 pathology among young children and infants was typically less severe compared to that observed among adults, this has not remained entirely consistent as SARS-CoV-2 variants have emerged. There is an enormous body of evidence demonstrating the benefits of human milk antibodies (Abs) in protecting infants against a wide range of enteric and respiratory infections. It is highly plausible that the same holds true for protection against SARS-CoV-2, as this virus infects cells of the gastrointestinal and respiratory mucosae. Understanding the durability of a human milk Ab response over time after infection is critical. Previously, we examined the Abs present in milk of those recently infected with SARS-CoV-2, and concluded that the response was secretory IgA (sIgA)-dominant and that these titers were highly correlated with neutralization potency. The present study aimed to monitor the durability of the SARS-CoV-2 IgA and secretory Ab (sAb) response in milk from COVID-19-recovered lactating individuals over 12 months, in the absence of vaccination or re-infection. This analysis revealed a robust and durable Spike-specific milk sIgA response, that at 9-12 months after infection, 88% of the samples exhibited titers above the positive cutoff for IgA and 94% were above cutoff for sAb. Fifty percent of participants exhibited less than a 2-fold reduction of Spike-specific IgA through 12 months. A strong significant positive correlation between IgA and sAb against Spike persisted throughout the study period. Nucleocapsid-specific Abs were also assessed, which revealed significant background or cross reactivity of milk IgA against this immunogen, as well as limited/inconsistent durability compared to Spike titers. These data suggests that lactating individuals are likely to continue producing Spike-specific Abs in their milk for 1 year or more, which may provide critical passive immunity to infants against SARS-CoV-2 throughout the lactation period.

## Introduction

As of March 2023, there have been over 760 million confirmed cases of severe acute respiratory syndrome coronavirus 2 (SARS-CoV-2) infection, causing 6.8 million deaths (1). Although in the early pandemic period, COVID-19 pathology among young children and infants was typically less severe compared to that observed among adults, this has not remained entirely consistent as SARS-CoV-2 variants have emerged. For example, hospitalization rates among children <4 years old increased 5x during the USA Omicron (B.1.1.529) variant wave (December 19, 2021-Feburary 19, 2022) compared to during the Delta variant period (B.1.617.2, June 27-December 18, 2021). During this Omicron wave, infants <6 months accounted for 44% of all COVID-19-related hospitalizations, and 21% required ICU admission (2). Especially as variants emerge that are more transmissible, children are at similar risk of SARS-CoV-2 infection as adults (3), and during the 2021-2022 waves of infections in the USA (*largely comprised of Delta (B.1.617.2) and Omicron (B.1.1.529) variants, respectively*), new cases in children aged 0-4 surpassed those among adults 65+, an epidemiological feature not previously observed (4). Notably, even asymptomatic infection can lead to ‘Multisystem Inflammatory Syndrome in Children’ (MIS-C), a rare but potentially deadly inflammatory condition, and young children are responsible for a significant amount of SARS-CoV-2 dissemination (5, 6). Clearly, protecting this population from infection remains essential, (7–9).

COVID-19 vaccines are available for infants >6 months, but those <6 months are not eligible. Although this is in part due to the potential for infants to receive vaccine- or infection-induced maternal IgG in utero, optimal placental IgG transfer requires timely maternal infection or vaccination, as well as surmounting the significant vaccine hesitancy among pregnant people (6). Importantly, maternal antibodies (Abs) wane from birth until they are undetectable by 6-12 months, with the kinetics of this decline being highly dependent on the initial titers transferred (10). Even for infants receiving high titers of maternal IgG in utero, milk Ab can provide critical complementary mucosal protection (11, 12). As such, the passive immunity of the Abs provided through human milk-feeding remains a highly relevant mechanism to protect infants from SARS-CoV-2.

Human milk contains approximately 0.7g/L IgA, which comprises ∼90% of the total immunoglobulin (Ig) in milk (13, 14). Approximately 2% of Ig in milk is IgG, as humans rely on placentally-transferred IgG for systemic immunity (14). Milk-derived Abs are likely most active in the oral/nasal cavity and upper gastrointestinal (GI) tract of infants, though ∼30% - 50% of these Abs have been shown to resist degradation in the stomach for as long as 2h suggesting that they are functional throughout the GI tract and possibly beyond (15). Milk IgG is derived primarily from the serum with a minority arising from local mammary production. B cells in the mammary gland that ultimately produce IgA (and to a lesser extent, IgM) that becomes secretory (s)IgA predominately originate from the gut-associated lymphoid tissue (GALT), exemplifying a critical *entero-mammary* link, wherein the sAbs found in human milk echo the immunogens identified in the maternal GI tract (and airways) (16, 17). This IgA is polymerized (mostly dimerized) with a joining (J) chain within the B cell prior to secretion, and then bound by the polymeric Ig receptor (pIgR) on mammary epithelial cells. PIgR is cleaved as it transports Ab into the milk, leaving the secretory component (SC) attached and resulting in sAb (16, 18). Determining whether or not sAbs are elicited in milk after infection or vaccination is critical, as this Ab class is highly stable and resistant to enzymatic degradation in milk and all mucosae - not only in the infant oral/nasal cavity, but in the airways and GI tract as well (14, 19).

Previously, we examined the Abs present in milk of those recently infected with SARS-CoV-2, and concluded that the response was sIgA-dominant and that sIgA titer was highly correlated with neutralization potency (20, 21). In the present study, milk samples from 16 COVID-19-recovered study participants were collected longitudinally for up to 12 months post-infection, in order to determine the durability of the sIgA response in milk over time. Specific reactivity against SARS-CoV-2 Spike and Nucleocapsid were measured.

## Material and methods

### Study participants and milk sampling

This study was approved by the Institutional Review Board (IRB) at Mount Sinai Hospital (IRB 19-01243). Individuals were eligible to have their milk samples included in this analysis if they were lactating and had a confirmed SARS-CoV-2 infection (by an FDA-approved COVID-19 PCR test) 3-8 weeks prior to the initial milk sample used for analysis as well as 9 months after infection. Participants were excluded if they had any acute or chronic health conditions affecting the immune system. Participants were recruited nationally via social media in April-June of 2020 and subject to an informed consent process. Certain participants contributed milk they had previously frozen for personal reasons, while most pumped samples specifically for this research project. All participants were either asymptomatic or experienced mild-moderate symptoms of COVID-19 that were managed at home. All demographic information on participant milk samples is shown in Table 1. Participants were asked to collect approximately 30mL of milk per sample into a clean container using electronic or manual pumps, and if able and willing, to continue to pump and save monthly milk samples. Milk samples were frozen at −20°C in participants’ home until samples were picked up and stored at −80°C until testing. Sample IDs are not known to anyone outside the research group.

**Table 1:**
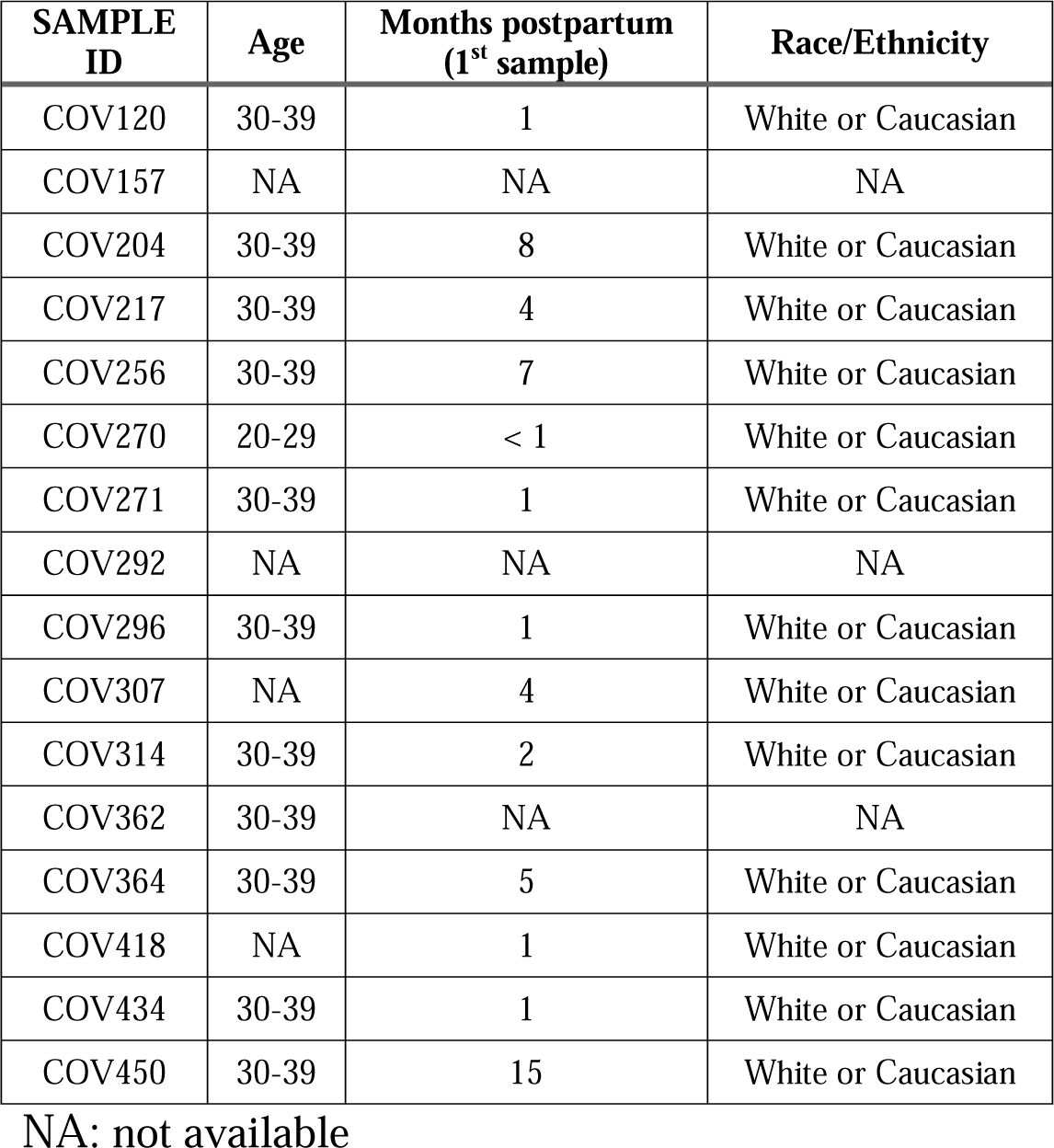
Participant Information

| **SAMPLE ID** | **Age** | **Months postpartum (1<sup>st</sup> sample)** | **Race/Ethnicity** |
| --- | --- | --- | --- |
| COV120 | 30-39 | 1 | White or Caucasian |
| COV157 | NA | NA | NA |
| COV204 | 30-39 | 8 | White or Caucasian |
| COV217 | 30-39 | 4 | White or Caucasian |
| COV256 | 30-39 | 7 | White or Caucasian |
| COV270 | 20-29 | < 1 | White or Caucasian |
| COV271 | 30-39 | 1 | White or Caucasian |
| COV292 | NA | NA | NA |
| COV296 | 30-39 | 1 | White or Caucasian |
| COV307 | NA | 4 | White or Caucasian |
| COV314 | 30-39 | 2 | White or Caucasian |
| COV362 | 30-39 | NA | NA |
| COV364 | 30-39 | 5 | White or Caucasian |
| COV418 | NA | 1 | White or Caucasian |
| COV434 | 30-39 | 1 | White or Caucasian |
| COV450 | 30-39 | 15 | White or Caucasian |

NA: not available

### Luminex assay

Milk samples were thawed, centrifuged at 800g for 15 min at room temperature, fat was removed, and the de-fatted milk transferred to a new tube. Centrifugation was repeated 2x to ensure removal of all cells and fat. Skimmed acellular milk was aliquoted and frozen at −80°C until use. SARS-CoV-2 Ab levels were measured via optimized Luminex assay. SARS-CoV-2 spike trimer (wild-type (WT) Wuhan-1 strain, gifted from the Florian Kramer lab) and Nucleocapsid protein (WT, ProSci) were coupled to the beads using xMAP Ab coupling kit according to manufacturer’s instructions (Luminex). SARS-CoV-2 Spike was coupled to the xMAP beads at 2.5 μg per million beads, and Nucleocapsid was coupled at 0.5 μg per million beads to account for differences in protein size. The coupled beads were stored in PBS solution with 0.5% Casein (Thermo Scientific) and 0.05% sodium azide (Bicca) at 4°C until use.

Milk samples were serially diluted from neat to 1:2187 with 0.5% casein buffer in a 96-well sample plate. 50 μL of milk was then transferred to a black 96 well microplate (Greiner). Coupled xMAP beads were then added to each well at 2500 beads/region in a total of 50 μL solution. The plate was covered with a black lid and shaken at room temperature for 60 min at 600 rpm. The beads were then captured by the magnetic plate holder and washed twice with 0.5% casein. 100 μL of 2 μg/ml of Biotin labeled goat anti-Human IgA (Thermo Fisher) or 6 μg/ml of Biotin labeled sheep anti-Human SC (Nordic-mubio) were added to the plates and shaken for 30 min at 600 rpm. Beads were washed twice after removing the supernatant, followed by adding 100 μL of Streptavidin-PE (BioLegend) at 1 μg/ml and shaken for 30 min. Beads were washed 2x and resuspended in 100 μL of casein buffer, and read by a Luminex FlexMAP3D device with xPONENT 4.2 software. Median fluorescent intensity (MFI) was measured for each well and exported for analysis.

### Analytical Methods

Control milk samples obtained prior to December 2019 were used to establish positive cutoff values for each assay. Milk was defined as positive at an optimized screening dilution of 1:18 if the sample exhibited an MFI 3 standard deviations (SD) above the mean MFIs of COVID-naïve control samples. Endpoint binding titers were calculated from log-transformed titration curves using 4-parameter non-linear regression, using MFI cutoff values of 650 and 150 for the IgA and sAb assays, respectively. Significance of longitudinal changes in Ab titers were analyzed by paired one-way ANOVA test. Correlation analyses were performed using the Spearman correlation test.

### Neutralization Assay

A proxy neutralization assay that detects Abs specific for the Spike receptor binding domain (RBD) that can prevent RBD-ACE2 binding was performed using milk samples, following manufacturer’s protocol (Genscript). Results were read on a Powerwave plate reader, and the percentage of virus neutralization was determined compared to the provided positive and negative controls.

## Results

### Longitudinal Spike-specific Ab profiles in milk after SARS-CoV-2 infection

Sixteen sets of milk samples obtained at sequential time points from COVID-19-recovered lactating individuals were examined (Table 1). Samples were analyzed for IgA and sAb reactivities against SARS-CoV-2 Spike and Nucleocapsid antigens using a Luminex-based immunoassay as described in methods. All milk samples obtained from COVID-19-recovered participants exhibited positive MFIs indicating Spike-specific IgA and sAb 3-8 weeks post infection (Fig. 1a, b). Longitudinal milk samples were similarly assessed, and endpoint titers determined for each assay. It was found that at the last time point available (9-12 months) 14/16 participants (88%) continued to exhibit positive IgA endpoint titers against Spike (Fig. 1c), and 15/16 participants (94%) continued to exhibit positive sAb endpoint titers against Spike (Fig. 1d).

**Figure 1.**
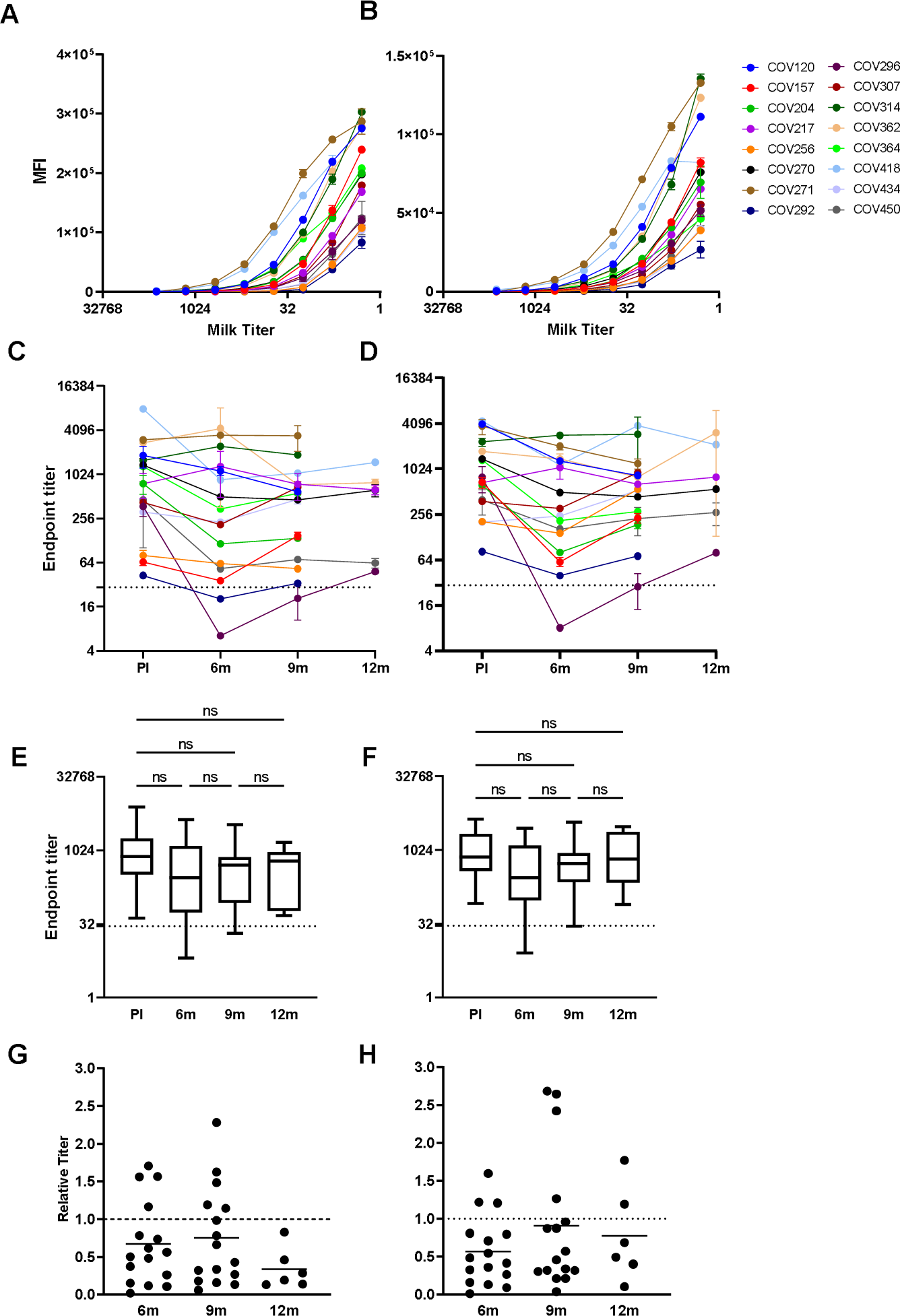
Longitudinal human milk IgA and sAb reactivity against SARS-CoV-2 Spike demonstrates significant durability over time. SARS-CoV-2 Ab levels were measured via optimized Luminex assay as described in methods. (A-B) Titration curves of IgA and sAb 3-8 weeks post-infection. A) IgA, B) sAb. (C-D) Individual endpoint titers over time. C) IgA, D) sAb. Dotted lines: positive cutoff value. (E-F) Grouped endpoint titers with mean values and ANOVA analysis for change over time. E) IgA, F) sAb. Dotted lines: positive cutoff value. (G-H) Relative endpoint titers over time compared to 3-8 weeks post-infection. G) IgA; H) sAb. Dotted line indicates relative titer of 1, meaning no change. PI: post-infection.

Endpoint titers were grouped by time point in order to determine if significant changes had occurred over the study period (Fig. 1e, f). Values were compared by paired ANOVA test. At the initial 3-8 weeks post-infection time point, samples exhibited a mean IgA endpoint titer against Spike of 1354 (range 43-3465). At the following time point examined, 5-6 months post-infection, samples exhibited a mean IgA endpoint titer against Spike of 960 (range 20-4323), which was not significantly different from the initial time point (Fig. 1e). At 9 months samples exhibited a mean IgA endpoint titer of Spike of 740 (range 33-3437), which was also not significantly different compared to either the initial time point or the 5-6 month time point. Following a highly similar trend, mean sAb titers against Spike at 3-8 weeks (mean 1436, range 82-4000), 5-6 months (mean 721, range 8-2835), and 9-12 months (mean 1007, range 28-3816) were not significantly different (Fig. 1f).

To evaluate the relative change of IgA and sAb levels over time, endpoint titers for each longitudinal milk sample were normalized by the individual’s endpoint titer of their initial milk sample obtained 3-8 weeks post infection (Fig. 1g,h). Notably, 94% of the participants exhibited < 10-fold reduction in IgA endpoint titer 5-6 months post infection, and 56% exhibited <2-fold reduction. By 9 months, 50% of participants exhibited <2-fold decrease in IgA titer, having decreased by only 25% on average compared to those at 3-8 weeks (range: 73% decrease to 127% increase). By the end of the study period, IgA had decreased by 64% (17% to 87%), however, this was not significantly different from the 9 month time point (Fig. 1g).

Examining the durability of Spike-specific sAb, COV296 and COV157 exhibited >10-fold decrease in titer at 5-6 months post infection, with the remaining participants exhibiting <10-fold decreases (titers were 57% of those compared to those observed at 3-8 weeks). At 9 months post infection, sAb titers were on average maintained at 90% of those measured at 3-8 weeks post infection, with only COV295 exhibiting >10-fold decrease, though this titer was also similarly increased compared to that measured at 5-6 months. Fifty percent of participants exhibited <2-fold reduction by the end of the study period (Fig. 1h).

### Longitudinal Nucleocapsid-specific Ab profiles in milk after SARS-CoV-2 infection

Similar analysis of IgA and sAb reactivity against SARS-CoV-2 Nucleocapsid protein was performed (Fig. 2). Despite the significant difference in IgA reactivity against Nucleocapsid exhibited by milk obtained from COVID-recovered donors compared to the COVID-naïve controls (p<0.0001), a positive MFI cutoff value could not be determined due to high background reactivity of the COVID-naïve control samples (Fig.2c, dashed line: mean naïve MFI + 3SD). Endpoint titers were calculated as above. At the initial 3-8 weeks post-infection time point, samples exhibited a mean IgA endpoint titer against Nucleocapsid of 649 (range 1-1098). By 5-6 months post-infection, samples exhibited a mean IgA endpoint titer against Nucleocapsid of 233 (range 2-1175), which was significantly decreased from the initial time point (Fig. 2, p=0.0066). By 9 months, samples exhibited a mean IgA endpoint titer against Nucleocapsid of 383 (range 4-1042), which was not significantly different compared to either the initial time point or the 5-6 month time point. The mean sAb titers against Nucleocapsid at 3-8 weeks (mean 981, range 13-4100), 5-6 months (mean 427, range 18-2450), and 9-12 months (mean 590, range 23-2150) were not significantly different (Fig. 2f).

**Figure 2.**
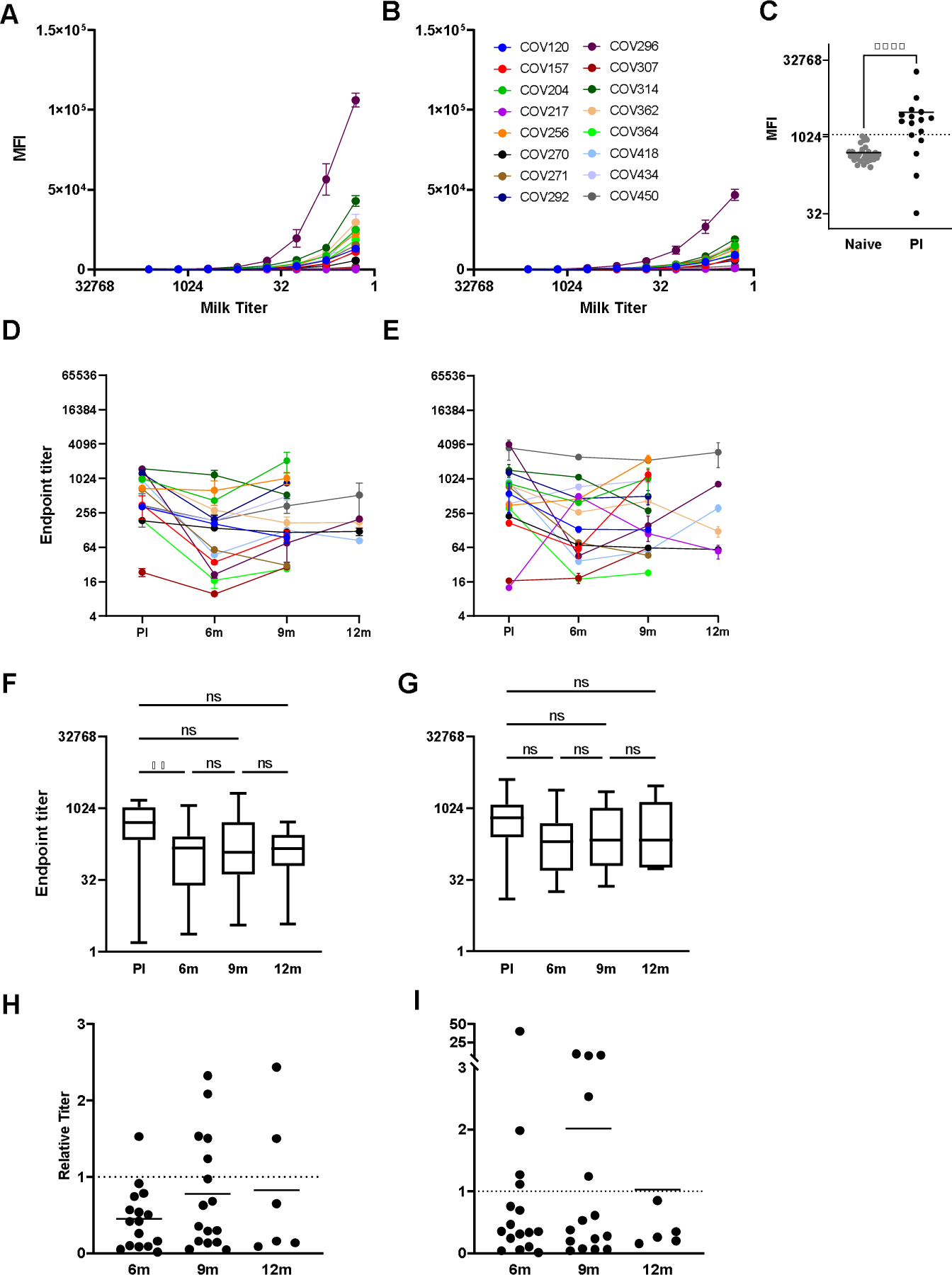
Longitudinal human milk IgA and sAb against SARS-CoV-2 Nucleocapsid are maintained less consistently compared to Spike Abs and subject to significant background reactivity. (A-C) Titration curves of IgA and sAb 3-8 weeks post-infection. A) IgA; B) sAb, C) high background IgA of COVID-19-naïve milk hinders determination of a positive cutoff value. MFI at 1/18 dilution (screening dilution) are shown. (D-E) Individual endpoint titers over time. D) IgA, E) sAb. (F-G) Grouped endpoint titers with mean values and ANOVA analysis for change over time. F) IgA, G) sAb. Relative endpoint titers over time compared to 3-8 weeks post-infection. H) IgA, I) sAb. Dotted line indicates relative titer of 1, meaning no change. PI: post-infection.

In contrast to the Spike data detailed above, the longitudinal changes observed for sAb titers (Fig. 2c, d) were inconsistent with changes observed for IgA. Specifically, COV217, COV314, COV362, and COV418 exhibited divergent IgA versus sAb titer changes over time. Compared to 3-8 weeks post-infection, the IgA titers decreased by an average of 22% at 9 months, while sAb titers increased by an average of 100% (Fig. 2e, f). ANOVA analysis determined there to be no significant changes in IgA or sAb titers across most time points, with the exception of a significant IgA decrease from 3-8 weeks to 5-6 months post-infection.

IgA and sAb titers against Spike and Nucleocapsid at each time point were compared by Spearman correlation analysis (Fig. 3). As shown in our previous work, a strong significant positive correlation was found between IgA and sAb Spike titers 3-8 weeks post infection (Fig. 3a, r=0.8765; p<0.0001). The correlation between IgA and sAb against Spike was maintained throughout the study period (Fig. 3c, r=0.9130, p<0.0001). In contrast, the correlation between IgA and sAb titers against Nucleocapsid at 3-8 weeks post-infection was only moderate (Fig. 3b, r=5747; P=0.0199), and no correlation was found in the samples 5-6 months post infection or beyond (data not shown). No correlation was found between anti-Spike and anti-Nucleocapsid IgA or sAb titers (Fig. 3d, e).

**Figure 3.**
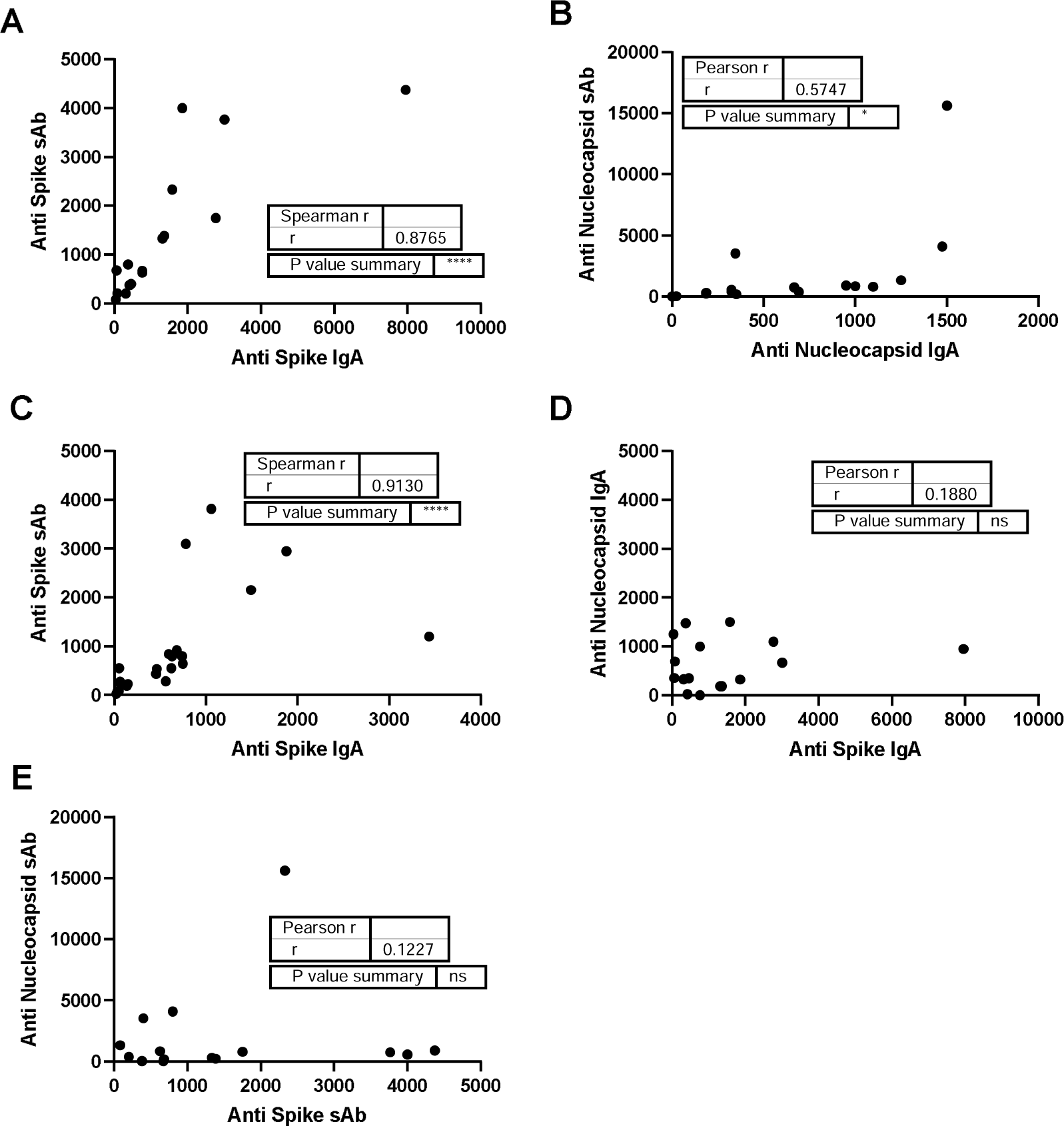
Spike-specific human milk IgA and sAb remain highly correlated throughout the study period, while Nucleocapsid Abs exhibit moderate correlation and Spike vs Nucleocapsid Ab reactivities are unrelated. A) Spike IgA vs sAb 3-8 weeks post-infection. B) Nucleocapsid IgA vs. sAb 3-8 weeks post-infection. C) Correlation between Spike IgA and sAb at the end of the study period. D) Spike vs Nucleocapsid IgA 3-8 weeks post infection. E) Spike vs Nucleocapsid sAb 3-8 weeks post infection. Endpoint titers were compared by Spearman correlation analysis.

### Durability of SARS-CoV-2 neutralization capacity of milk from COVID-19-recovered donors over time

A proxy neutralization assay that measures Abs specific for the Spike receptor binding domain (RBD) that block the RBD-ACE2 interaction was performed using a premade kit (Genscript). Eleven pairs of samples analyzed above were examined for changes in neutralization capacity from 3-8 weeks post-infection to 9 months post-infection. At 3-8 weeks post-infection, the mean % neutralization of milk at neat was 49% (range 30% - 73%), with all samples exhibiting neutralization activity that was significantly greater than that exhibited by naïve milk (p=0.0029). At 9 months, the mean % neutralization was 47% (range 30% - 67%) (Fig. 4). Although one sample’s neutralization capacity dropped below that of the naïve controls at 9 months (COV364, from 63% to 19%). Notably, COV364 dropped 67% in IgA titer and 89% in sAb titer at 9 months post infection. It was found that no significant change in neutralization was detected between the early and late groups.

**Figure 4.**
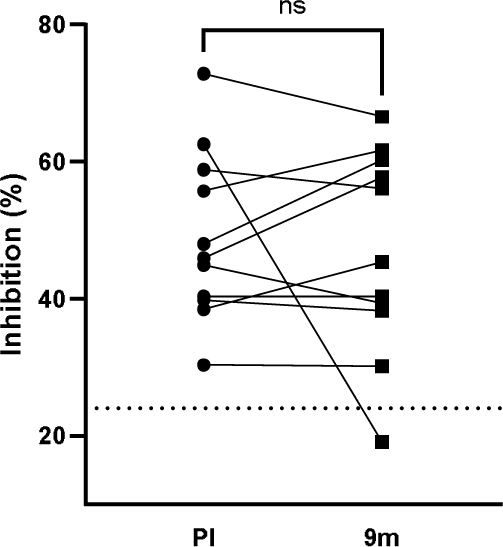
SARS-CoV-2-neutralizing Ab in human milk is maintained for at least 9 months after infection. A proxy neutralization assay that measures Abs specific for the Spike receptor binding domain (RBD) that block the RBD-ACE2 interaction was performed using a premade kit according to instructions (Genscript). Dotted line: background neutralization level of COVID-19-naïve milk. Paired t-test was used for analysis. PI: post-infection.

## Discussion

Nearly all sAb derives from the GALT, via the *entero-mammary* link *-* homing of Ab-secreting B cells from the gut to the mammary gland. Various animal studies have demonstrated this migration and homing during late pregnancy and lactation. Homing appears to be controlled hormonally, as well as by various adhesion factors on the B cells and the maternal vasculature including MadCAM-1, integrin α4β7, CCL28, and CCR10 (22). This link is an evolutionarily critical mechanism facilitating specific protection to a vulnerable infant against the pathogens in the maternal/infant environment sampled by the maternal GALT, and provides key immunological training for the infant (22). Infants benefit greatly from the sAb provided in human milk, as the neonate mucosal immune system is relatively deficient in sAb production as well as other key immune factors. Even past the neonatal period, these Abs can supplement the infant’s own immunity to provide protection against pathogens against which the infant does not yet have immunological protection. sAbs are critical due to their relative durability in harsh environments including the milk itself, and all mucosal fluids including the oral/nasal cavity and GI tract (14, 15).

SARS-CoV-2 does not transmit via human milk. Though viral RNA has been identified in a small minority of milk samples studied (23–25), infectious SARS-CoV-2 particles have not. Several meta-analyses to date have failed to find evidence of worsened health outcomes for infants of any age chest/breast-fed by a SARS-CoV-2-infected person, or increased rates of infection for these infants compared to those who are formula-fed (26–28). We and others have reported SARS-CoV-2-specific Abs in milk obtained from donors with previous infection (21, 29–31). Our prior work determined that infection elicits a robust Spike-specific milk IgA and sAb response in ∼ 90% of cases that is highly neutralizing. In the present study, we aimed to monitor the SARS-CoV-2 IgA and sAb response in the milk from COVID-19-recovered individuals longitudinally over 1 year. Unlike the observed decay over time of the serum response that has been widely cited in other reports(32–35), we observed little change in Spike-specific milk titers over time. Notably, 50% of participants exhibited <2-fold reduction of Spike-specific IgA, and 94% of participants sustained positive sAb titers against Spike through 12 months. IgA and sAb titers maintained a strong significant positive correlation throughout the study period. Abs against Nucleocapsid protein have not been intensively studied because these Abs would be non-neutralizing; however, these Abs are useful as an indicator of pre-vaccine or breakthrough infections, and likely play a key role in clearing virus-infected cells or modulating immune response (36). As such, the longevity of the Nucleocapsid Ab response remains highly informative and worth investigation. Previous studies have demonstrated cross-reactivities among CoV-229E, CoV-OC43, MERS, SARS Nucleocapsid in the sera of COVID-19 recovered patients (37), which may explain the high level of background observed among the COVID-naïve samples, as well as the erratic titers measured over time relative to those against Spike.

The serological response against SARS-CoV-2 varies by variant, disease severity, and pre-existing conditions (38). Generally, the decay rate of Spike/RBD-specific serum IgA is high. The half-life of RBD-specific IgA has been found to be ∼71 days, with certain reports finding serum IgA levels to be undetectable after 60 days (32, 39, 40). Serum IgG half-life has been reported in multiple studies to be >3 months, with 57% of individuals who were asymptomatically infected to be IgG seropositive after 8 months, with 71% of those who experienced mild symptoms maintaining positive IgG titers (32, 39, 40). In the present study, Spike-specific sIgA titers in milk were shown to be significantly more durable over time.

The highly durable titers observed in the present study may be reflective of long-lived plasma cells in the GALT and/or mammary gland, as well as continued antigen stimulation in these compartments, possibly by other human coronaviruses, or repeated exposures to SARS-CoV-2. Consistent with the present data, it has been shown in general that although an initial IgA plasmablast response quickly declines, IgA-producing plasma cells can persist for decades in the human GALT, even when not measurable in the periphery (41). Given the established entero-mammary link, the highly durable Spike-specific sIgA in milk observed herein strongly suggests similarly long-lived plasma cells in the mammary gland.

The present data suggest that lactating individuals who recover from COVID-19 are likely to continue producing specific sIgA in their milk for 1 year or more. Based on studies of other mucosal pathogens, milk sIgA likely provides critical passive immunity to infants against SARS-CoV-2 infection and COVID-19 pathology. The significant durability of the Spike-specific sIgA response in milk highlights the importance of continued chest/breast-feeding after SARS-CoV-2 infection. Given the present lack of knowledge concerning the potency, function, durability, and variation of the human milk immune response not only to SARS-CoV-2 infection, but across this understudied field in general, the present data contributes greatly to filling immense knowledge gaps and furthers our work towards *in vivo* efficacy testing of extracted milk Ab in the COVID-19 pandemic context and beyond.

## Data Availability

All data produced in the present study are available upon reasonable request to the authors

## Acknowledgements

As always, we are indebted to the milk donors who make this work possible. Spike protein was generously gifted from the Krammer lab. This study was funded by the NIH/NIAID grant number R01 AI158214.

